# Adverse childhood experiences and their impacts on subsequent depression and cognitive impairment in Chinese adults

**DOI:** 10.1101/2022.09.07.22279699

**Authors:** Tiantian Zhang, Lena Kan, Changbo Jin, Wenming Shi

**Affiliations:** School of Social Development and Public Policy, Fudan University, Shanghai, China; Department of International Health, Bloomberg School of Public Health, Johns Hopkins University, Baltimore, Maryland, USA; Tongji University School of Medicine, Shanghai, China; School of Public Health, Fudan University, Shanghai, China

**Keywords:** Adverse childhood experiences, Depression, Cognitive impairment, Physical abuse, Adults

## Abstract

**Background:** Adverse childhood experiences (ACEs) are prevalent and have long lasting effects. This study aimed to explore the associations between ACEs exposure with subsequent depression and cognitive impairment and to assess whether sociodemographic characteristics modify these associations.

**Method:** A total of 14,484 participants from the China Health and Retirement Longitudinal Study (CHARLS) 2015 and life history survey in 2014 were enrolled. Depression was assessed by the 10-item Center for Epidemiologic Studies Depression scale. Cognitive performance was evaluated by three composite measures: episodic memory, mental intactness and global cognition. A wide range of 12 ACE indicators were measured by a validated questionnaire. Multiple regression models and stratified analysis explore the relationship between accumulated ACEs with subsequent depression and cognitive impairment and potential modifiers.

**Results:** Compared with individuals without ACEs, those who experienced four or more ACEs have a higher risk of subsequent depression (adjusted odds ratio, aOR=2.65, 95% confidence intervals [CIs]: 2.21-3.16), poorer mental intactness (β= -0.317 [-0.508 to -0.125]) and worse global cognition (β= -0.437 [-0.693 to -0.181]). Trend analyses showed a dose-response association between accumulated ACEs with subsequent depression and cognitive impairment. The modifications of the association by age, sex, educational level and family’s financial status during childhood were not observed.

**Conclusion:** Our study suggests that higher ACEs exposure increases the risk of subsequent depression and cognitive impairment in Chinese adults regardless of sociodemographic characteristics. The findings provide important implications for mitigating the adverse effects of early-life stress and promoting health in adulthood.

## Introduction

Adverse childhood experiences (ACEs), refer to a wide range of intensive stressors that occur before the age of 18, such as emotional neglect and physical abuse, which may affect children while growing up (Hughes et al., 2021). The theoretical framework from previous studies indicates that early-life trauma and adversities affect social, emotional and psychological domains through changes in neurodevelopmental trajectories (Shonkoff and Garner, 2012), and subsequently lead to an increased risk for lifelong health and well-being (Hughes et al., 2021).

Depression (depressive symptoms and depressive disorders) and cognitive impairment represent big challenges to global public health as both are highly prevalent (Kessler et al., 2009; Lu et al., 2021; Richardson et al., 2019). A growing number of studies indicated that accumulated ACEs were linked to alterations in brain structures and poorer neurodevelopmental functions across a variety of brain regions, such as limbic, hippocampal, prefrontal, and increased risks of depression, anxiety and suicide attempts later in life (Abbott and Slack, 2021; Dannlowski et al., 2012; Hawkins et al., 2021; Polanco-Roman et al., 2021). However, most previous studies are primarily from western countries including North America (Gould et al., 2012; Hawkins et al., 2021) and Europe (Houtepen et al., 2020), and few studies have focused on these associations in developing countries such as China, where ACEs are more prevalent (Houtepen et al., 2020).

The conventional ACE scale, which consists of 10 items determined by the Centers for Disease Control and Prevention (CDC) in the Kaiser study, is mostly used as an assessment tool in past publication (Felitti et al., 1998). Nevertheless, many concerns in recent years suggest that the conventional ACE scale may not adequately reflect the perceived childhood adversities in other populations (Cronholm et al., 2015). A set of new ACE indicators, such as unsafe neighbourhood, parental disability or sibling death have also been reported to be prevalent in ACE-related study (Cronholm et al., 2015; Rod et al., 2020). Socioeconomic and demographic characteristics, whether independent or clustered together, may be compounded to affect the association between ACEs exposure and health outcomes (Houtepen et al., 2020; Naicker et al., 2021). However, due to the differences in regional culture, study design and population, the findings are not consistent (Houtepen et al., 2020; Schickedanz et al., 2021). To build upon existing literature, a large-scale study with comprehensively clustering of ACE indicators is timely needed.

In this study, we used data from the China Health and Retirement Longitudinal Study (CHARLS), a national representative sample in China, to investigate the association of early-life exposure to accumulated ACEs and each individual indicator with depression and cognitive impairment in later life. Furthermore, we also evaluated whether the associations were modified by sociodemographic characteristics of age, sex, educational level and family’s financial status during childhood.

## Methods

### Study design and participants

This study was based on the CHARLS project, a nationwide representative survey of middle-aged and older adults covering 450 villages/resident committees from 28 provinces across mainland China. The details about the study design and sampling method have been reported in previous literature(Zhao et al., 2014). Briefly, the participants were recruited by a multistage probability sampling strategy, 17708 respondents were included in the baseline survey, and follow-up every two years. During the survey, the information on sociodemographic characteristics, behaviour and lifestyles, health conditions and functions as well as related biomarkers were collected. In the current study, we used the third wave of CHARLS study in 2015 consisting of 21,095 individuals, and then matched to the 2014 CHARLS life history survey (N=20,544). A total of 18,678 participants who completed both the two surveys were enrolled. After excluding individuals with missing data on mental health outcomes, ACE indicators and other possible confounders, a total of 14,484 participants from 126 Chinese cities were included in the final analysis. The geographical locations of the cities are displayed in Figure 1. Ethics approval was obtained from the Biomedical Ethics Review Committee of Peking University (Nu: IRB00001052-11015). All participants provided their written informed consents. The flowchart showed the participants’ enrollment (Figure S1).

**Figure 1.**
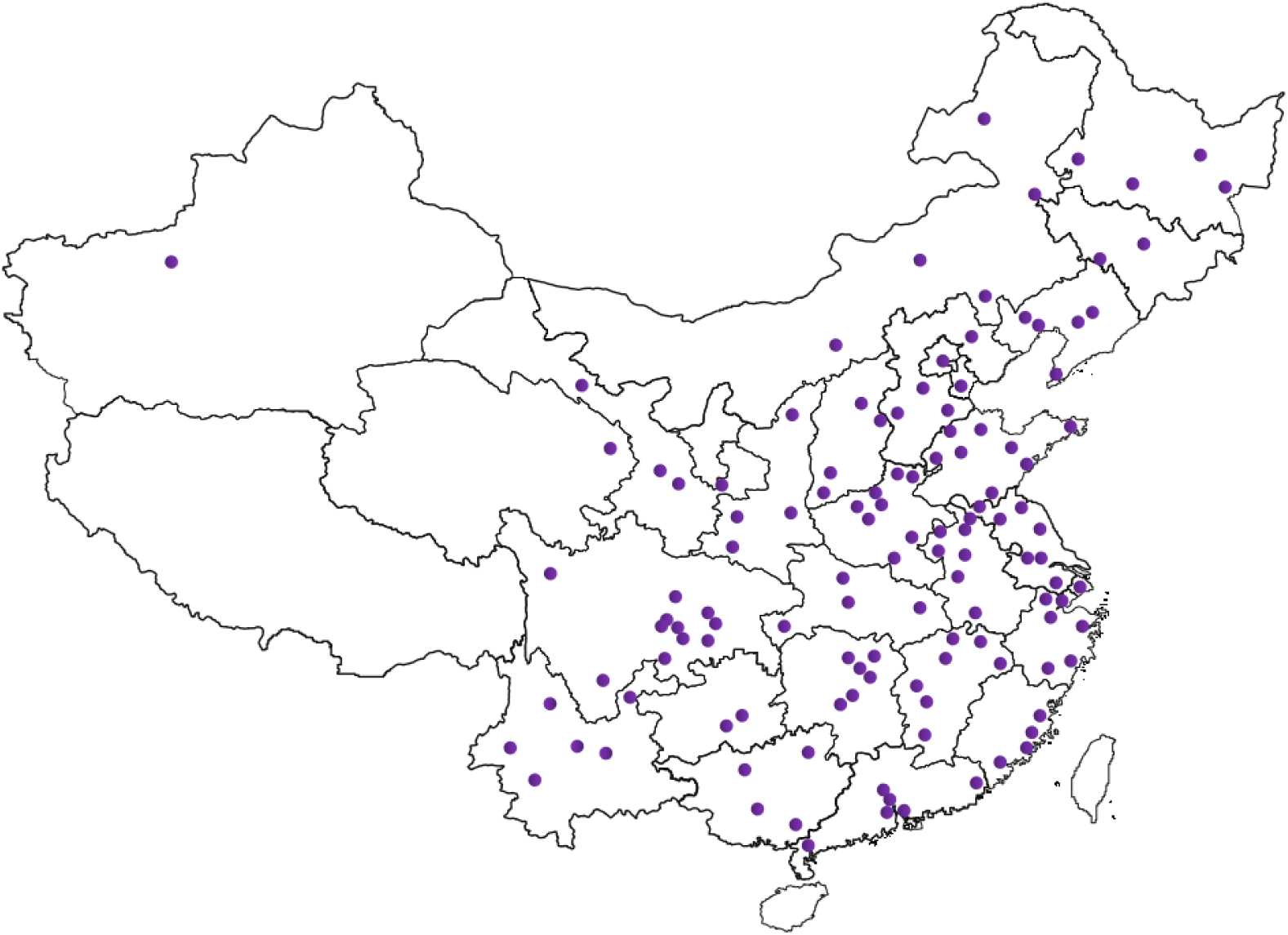
The geographical locations of the 126 cities in mainland China.

### Measurement of depression

Depression was measured by using the 10-item Center for Epidemiologic Studies Depression scale (CES-D-10). The validity of the CES-D-10 has been examined among Chinese adults (Boey, 1999). Each question was used to measure the frequency of a specific type of negative mood, where 0 indicated rarely or none; 1(some days); 2 (occasionally) or 3 (most of the times), respectively. We calculated the total of each specific-question score as the depressive score to assess the general status of depression. The total score ranges from 0 to 30, with a higher score indicating more depressive disorders. A categorical variable for the CES-D-10 score was applied to evaluate the depression based on a cut-off of 12(Li et al., 2019).

### Cognitive performance assessment

In line with previous CHARLS publications (Li et al., 2017; Rong et al., 2020), three composite measures of cognition score were adopted: a word-recall test, Telephone Interview of Cognitive Status (TICS-10) and a figure drawing test. The word-recall test was performed to assess the episodic memory, which asked the respondents to memorize and immediately recall ten Chinese nouns that had just been read to them in any order (immediate recall) and recalled the same word list after four minutes (delayed recall). The episodic memory score consists of the average of the immediate recall and delayed recalls, which ranged from 0 to 10. The TICS-10 and a figure drawing test were performed to evaluate the mental intactness related to orientation to time and attention, and visuospatial abilities. Ten mental status questions from the TICS including serial subtraction of 7 from 100 (up to five times), date (month, day, year), day of the week, and season of the year were recorded, and subjects were requested to redraw a picture of two overlapped pentagons. These scores were summed as a mental intactness indicator and ranged from 0 to 11. The global cognition score was generated by summing up the score of episodic memory and mental intactness, and considered as the primary outcomes of interests on cognitive performance. The global cognition score ranged from 0 to 21, with a higher score indicating a better cognition function (Rong et al., 2020).

### Measurement of adverse childhood experiences

We used a wide range of 12 ACE indicators from the CHALRS study, which could be classified into four categories: 1) child maltreatment (physical abuse, emotional neglect); 2) exposure to violence (domestic violence, peer bullying and unsafe neighbourhood); 3) parent/ sibling death or disability (parental death, parental disability, and sibling death); 4) parental maladjustment (household mental illness, substance abuse, parental separation or divorce and incarcerated household member). Table S1 presents the details about the definition and questionnaire descriptions of each items of individual indicator. Response to each ACE indicator was dichtomized (0 vs.1) and then summed to generate a cumulative number for each participant, ranging from 0 to 12. We classified the participants into five subgroups according to the cumulative numbers of ACE indicators: 0, 1, 2, 3 and ≥ 4.

### Covariates

Individual information on sociodemographic characteristics, behaviour and lifestyles, and self-reported chronic conditions were collected by a validated CHARLS questionnaire. The sociodemographic characteristics included age, sex, body mass index (BMI), educational level, marriage status (married v.s unmarried), maternal education, paternal education, and family’s financial status during childhood (which assessed by the question “When you were a child before the age of 17 years, compared to the average family in the same community/village, how was your family’s financial situation?” and was divided into two categories: same as or better than others vs. worse than others). The behaviour and lifestyles factors including cigarette smoking status (current; former: quitted smoking ≥ three years; never), alcohol consumption, social activity (yes vs. no) and night sleep duration were also recorded. The self-reported chronic diseases such as hypertension, diabetes and heart diseases were collected.

### Statistical analyses

Descriptive statistics were performed for all sociodemographic characteristics, behaviour and lifestyle and chronic conditions data. The prevalence of subsequent depression and average cognitive score were calculated and compared among each ACE subgroup, respectively. Multivariate logistics regression and generalized linear regression models were conducted to explore the relationship between each items of individual ACE indicator or accumulated ACEs and risk of depression and cognition disorders decline in later life, respectively. Both the crude and adjusted models were performed. The odds ratio (OR) or β-coefficient and 95% confidence intervals (95%CI) were calculated to report the association. The covariates in the adjusted model included age, sex, BMI, educational level, marriage status, maternal education, paternal education, family’s financial status during childhood, cigarette smoking status, alcohol consumption, social activity, night sleep duration and chronic diseases. Trend analyses were used to examine whether a dose-response association was present.

To examine the effect modifiers of several sociodemographic variables, stratified analyses were use by the following variables: sex (men compared with women), age (≥ 65 years compared with < 65 years), educational level (≥ middle school compared with < middle school) and family’s financial status during childhood (same as or better than others compared with worse than others). Interaction of accumulated ACEs and each aforementioned variable was also evaluated in the regression models.

In addition to the main analysis, we performed a sensitivity analysis by exploring the association between accumulated ACEs exposure of seven conventional indicators as reported in a previous publication (Lin et al., 2021) with depression and cognitive impairment in our study.

All statistical analyses were performed by STATA 16.0 (Stata Corp., College Station. TX, USA) and R software (version: 4.1.2). All *p* values were two-sided, and significance was defined as *p* < 0.05.

## Results

All the sociodemographic characteristics, behaviours and lifestyles factors, chronic diseases according to the number of ACEs were summarized (Table 1). Of the 14,484 participants enrolled, 6925 (47.8%) were men, and the average age was 60.7± 9.5 years. Overall, 79.7% of the participants reported experiencing one or more ACEs, and 10.5% had four or more ACEs (Table 1). Compared with those without any ACEs, individuals who experienced four or more ACEs were more likely to be older, of low educational level, unmarried, of worse financial status during childhood, and more likely to have less sleep and poorer health conditions (Table 1). The prevalence of each item of the ACE indicators as well as accumulated ACEs groups are shown in Figure 2. We observed that the prevalence of depression was associated with an increasing trend with the growing number of ACE indicators (Figure 3). Regarding cognitive performance, we found that the mean score of episodic memory, mental intactness, and global cognition were decreased along with an increasing number of ACEs (Figure 3).

**Table 1.** Descriptive characteristics of participants by number of ACEs^a^ [Mean ± SD or N (%)].

| Variables | ACEs (N=14484) |  |  |  |  | P-value |
| --- | --- | --- | --- | --- | --- | --- |
|  | 0<br>(N=2935) | 1<br>(N=4472) | 2<br>(N=3518) | 3<br>(N=2045) | ≥ 4<br>(N=1514) |  |
| Age (years) | 60.29±9.52 | 60.84±9.51 | 61.02±9.61 | 60.48±9.31 | 60.68±9.47 | 0.012 |
| Sex (%) |  |  |  |  |  |  |
| Men | 1296 (44.2) | 2116 (47.3) | 1743 (49.5) | 1027 (50.2) | 743 (49.1) | <0.001 |
| Women | 1639 (55.8) | 2356 (52.7) | 1775 (50.5) | 1018 (49.8) | 771 (50.9) |  |
| BMI (kg/m<sup>2</sup>) | 23.95±3.85 | 23.81±3.86 | 23.77±3.71 | 23.78±3.59 | 23.45±3.81 | 0.001 |
| Educational level |  |  |  |  |  |  |
| ≥ Middle school | 1142 (38.9) | 1572 (35.1) | 1089 (31.0) | 638 (31.2) | 424 (28.0) | <0.001 |
| < Middle school | 1793 (61.1) | 2900 (64.9) | 2429 (69.0) | 1407 (68.8) | 1090 (72.0) |  |
| Marriage status |  |  |  |  |  |  |
| Married | 2576 (87.8) | 3901(87.2) | 3033 (86.2) | 1771 (86.6) | 1271(83.9) | 0.006 |
| Unmarried | 359 (12.2) | 571 (12.8) | 485 (13.8) | 274 (13.4) | 243 (16.1) |  |
| Maternal education |  |  |  |  |  |  |
| ≥ Middle school | 1106 (37.7) | 1485 (33.2) | 1274 (36.2) | 804 (34.4) | 540 (35.6) | <0.001 |
| < Middle school | 1829 (62.3) | 2987 (66.8) | 2244 (63.8) | 1341 (65.6) | 974 (64.4) |  |
| Paternal education |  |  |  |  |  |  |
| ≥ Middle school | 378 (12.9) | 538 (12.0) | 404 (11.5) | 217 (10.6) | 166 (11.0) | 0.106 |
| < Middle school | 2557 (87.1) | 3934 (88.0) | 3114 (88.5) | 1828 (89.4) | 1348 (89.0) |  |
| Family's financial status during childhood |  |  |  |  |  |  |
| Same as or better than others | 2140 (72.9) | 2928 (65.5) | 2028 (57.6) | 991 (48.5) | 620 (41.0) | <0.001 |
| Worse than others | 795 (27.1) | 1544 (34.5) | 1490 (42.4) | 1054 (51.5) | 894 (59.0) |  |
| Cigarette smoking status |  |  |  |  |  |  |
| Current | 740 (25.2) | 1241(27.8) | 1042 (29.6) | 601 (29.4) | 457 (30.2) | 0.002 |
| Never | 1802 (61.4) | 2629 (58.8) | 1992 (56.6) | 1162 (56.8) | 859 (56.7) |  |
| Former | 393 (13.4) | 602 (13.4) | 484 (13.8) | 282 (13.8) | 198 (13.1) |  |
| Alcohol consumption |  |  |  |  |  |  |
| Yes | 909 (31.0) | 1490 (33.3) | 1309 (37.2) | 732 (35.8) | 593 (39.2) | <0.001 |
| No | 2026 (69.0) | 2482 (66.7) | 2209 (62.8) | 1313 (64.2) | 921 (60.8) |  |
| Social activity |  |  |  |  |  |  |
| Yes | 1570 (53.5) | 2457 (54.9) | 1877 (53.4) | 1114 (54.5) | 838 (55.4) | 0.484 |
| No | 1365 (46.5) | 2015 (45.1) | 1641 (46.6) | 931 (45.5) | 676 (44.6) |  |
| Night sleep duration (hours) | 6.61±1.88 | 6.47±1.92 | 6.29 ±1.94 | 6.19±2.03 | 5.89 ±2.12 | <0.001 |
| Chronic diseases |  |  |  |  |  |  |
| Hypertension | 688 (23.4) | 1054 (23.6) | 848 (24.1) | 478 (23.4) | 303 (20.0) | 0.031 |
| Diabetes | 172 (5.9) | 251 (5.6) | 197 (5.6) | 139 (6.7) | 98 (6.4) | 0.279 |
| Heart diseases | 351 (12.0) | 521 (11.6) | 448 (12.7) | 265 (13.0) | 192 (12.7) | 0.448 |

a: ACEs indicated adverse childhood experiences.

**Figure 2.**
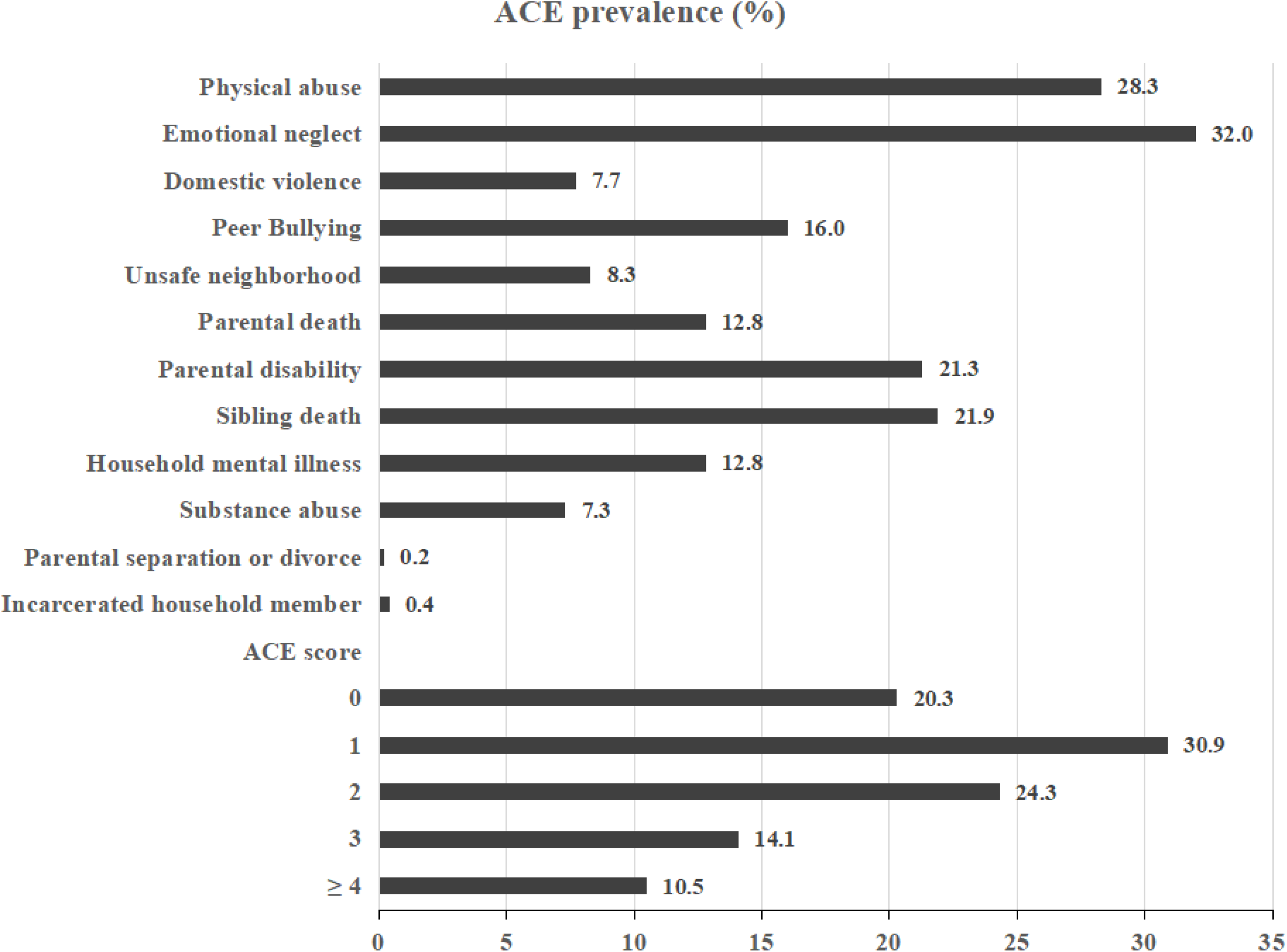
The prevalence of each item of ACEs among the participants.

**Figure 3.**
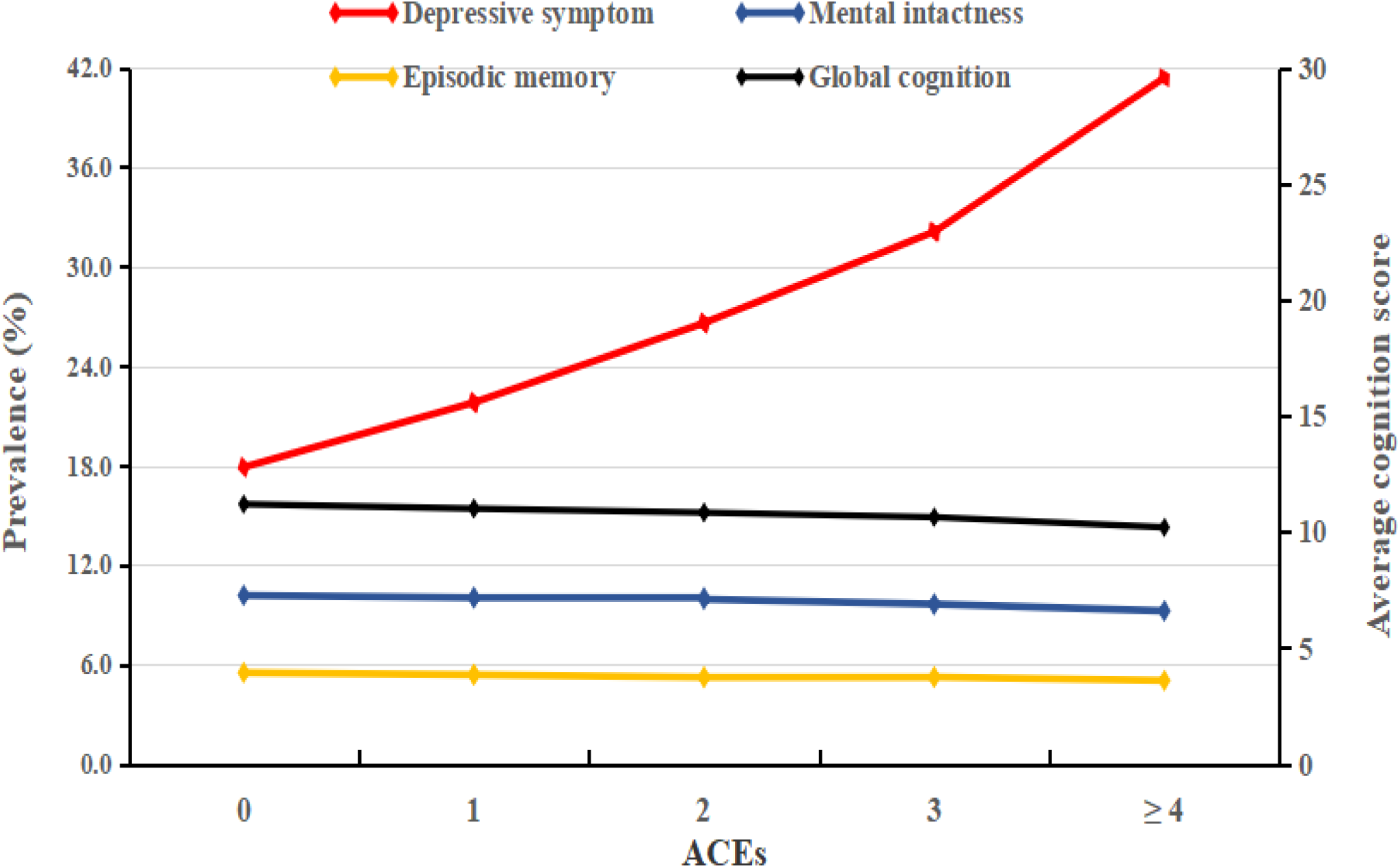
The trend of depression prevalence and average cognitive score according to the number of ACEs.

The multivariate regression model showed that individuals who experienced four or more ACEs had a significantly higher risk of depression and cognition decline later in life, compared with those without ACEs. After controlling for related confounders, the adjusted OR or β and 95%CI were 2.65 (2.21, 3.16) for depression, -0.317 (-0.508, -0.125) for mental intactness, and -0.437 (-0.693, -0.181) for global cognition score, respectively. The trend analysis indicated a significant dose-response relationship between cumulative ACEs exposure with risk of subsequent depression and cognitive impairment (P_trend_ ranged from 0.033 to <0.001, Table 2). Meanwhile, no significant association between accumulated ACEs exposure and episodic memory in adulthood was observed in model □ (Table 2). When looking at the specific item of ACE indicators, the results showed that both household mental illness and unsafe neighbourhood were significantly related with increased risks of depression and cognition decline. In addition, we also found that the risk of subsequent depression was significantly increased when early-life exposure to physical abuse (aOR:1.34, 1.20-1.48), domestic violence (aOR:1.44, 1.23-1.69), peer bullying (aOR:1.73, 1.53-1.96), parental disability (aOR:1.54, 1.39-1.72) or sibling death (aOR:1.21, 1.09-1.36) separately. Meanwhile, a significant association of cognitive disorders was observed in those exposure to specific individual indicator of substance abuse and parental death. Details of the association of each item of ACEs are shown in Figure S3. The stratified analysis presented similar outcomes and a significant dose-response relationship. However, we didn’t observe that sociodemographic characteristics of age, sex, educational level and family’s financial status during childhood significantly modified the association between ACEs and health outcomes among the participants (P all > 0.05, Table 3).

**Table 2.**
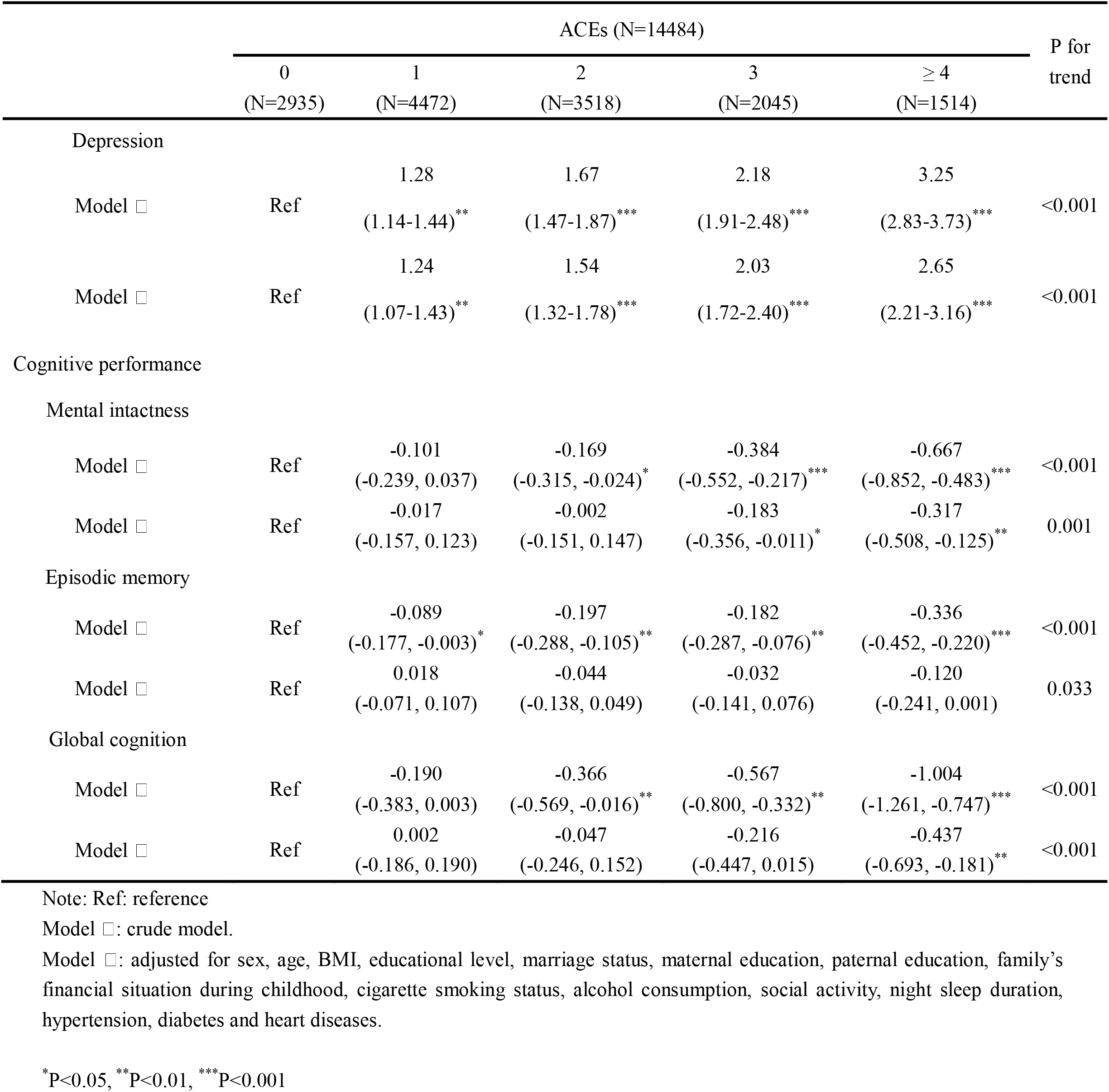
The association between adverse childhood experiences with depression and cognitive performance in adulthood.

**Table 3.**
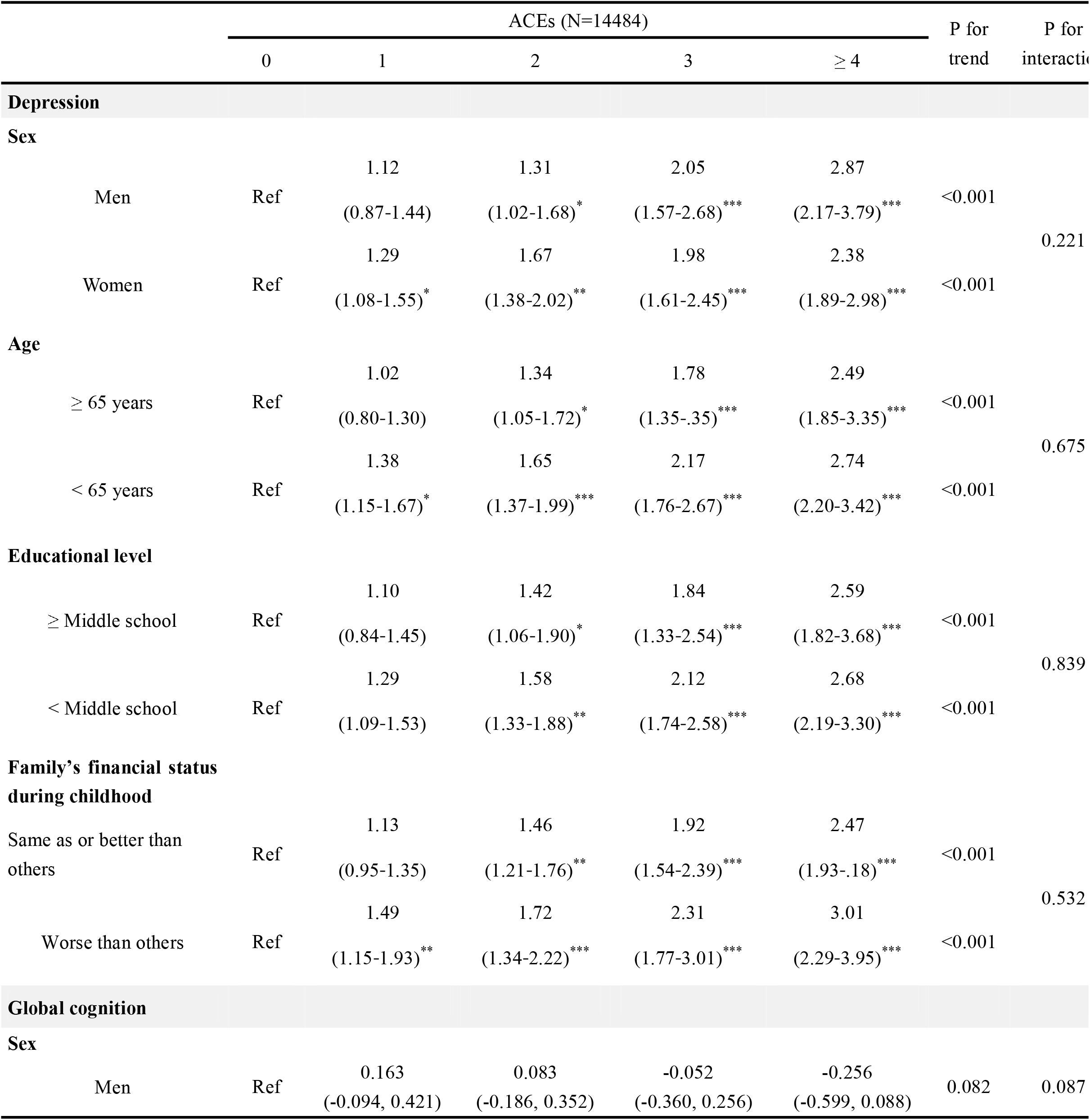
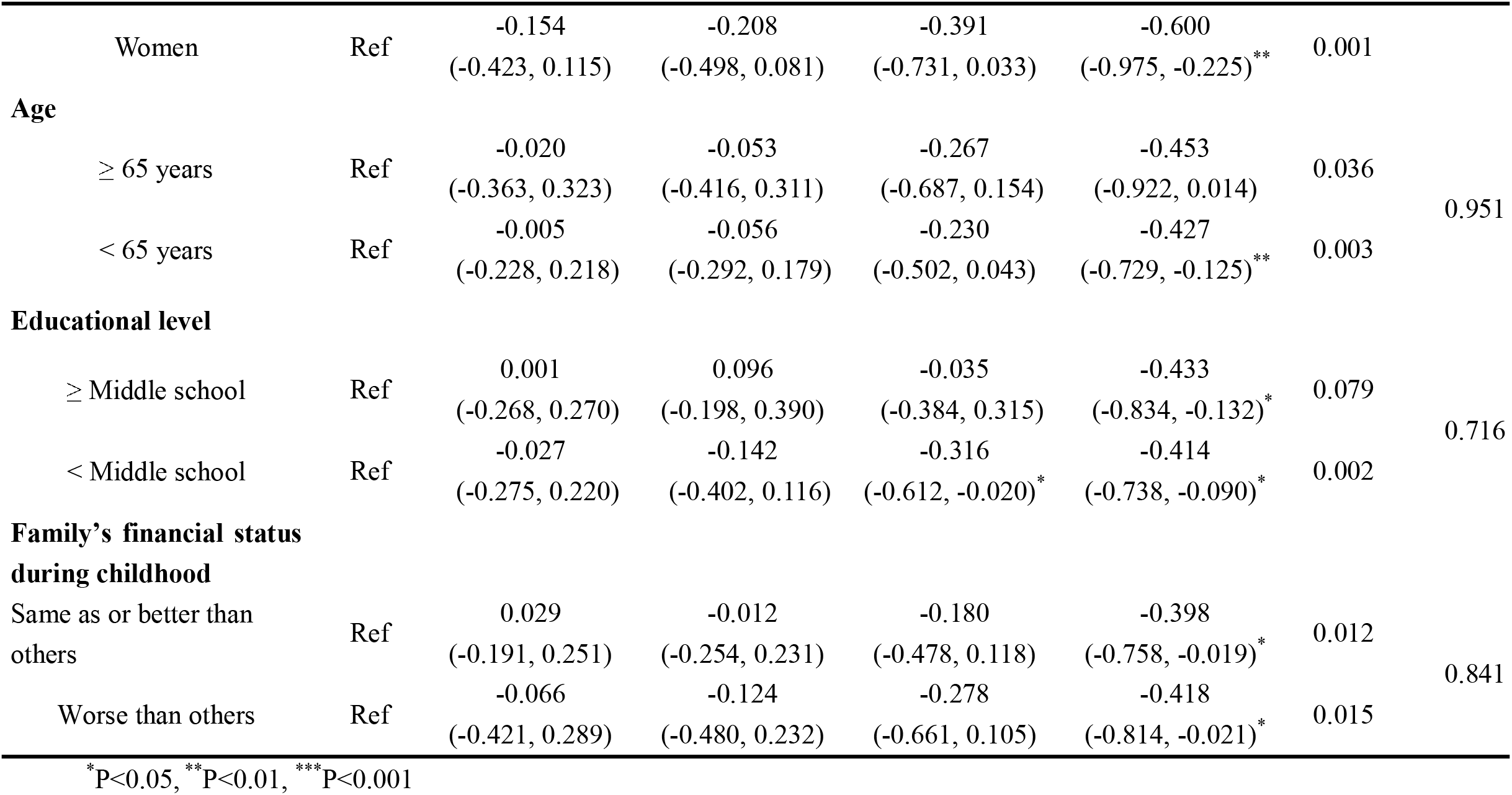
Subgroup analysis of ACEs exposure with subsequent depression and global cognition among the subjects.

Similar associations were also observed in the sensitivity analysis when exploring the seven conventional ACEs indicators exposure with depression and cognitive impairment (Table S3).

## Discussion

This nationwide population-based study suggests that exposure to ACEs was significantly associated with an increased risk of subsequent depression and cognitive impairment in Chinese adults. A significant dose-response relationship between the accumulated ACEs and subsequent depression and cognition decline was observed. Our findings did not show that sociodemographic characteristics of sex, age, educational level, and family’s financial status during childhood modify the association between early-life exposure to childhood advertises with mental health later in life.

Similar to previous studies in western countries (Abbott and Slack, 2021; Dantchev et al., 2019; Gould et al., 2012), we observed significant associations between exposure to ACEs with adult mental health, despite cultural differences. Given the insufficiency of conventional ACE scale from the CDC-Kaiser ACE study was generated based on a sample of mostly white and educated individuals(Felitti et al., 1998), we evaluated a more comprehensive range of ACE indicators in this study. Consistent with existing literature(Gal et al., 2011; Wang, 2020), our findings observed that physical abuse during early-life, domestic violence or peer bullying were related with an elevated risk of adult depression. For cognitive performance, we found that the specific categories of ACE indicators of parental maladjustment, i.e: household mental illness and substance abuse were linked to an elevated risk of cognitive impairment in adulthood, which was in parallel with previous studies (Bennett et al., 2012; Schickedanz et al., 2021). A likely explanation was that familial factors during childhood including shared genetic factors and a living environment have long-lasting effects on children’s health (Ferraro and Shippee, 2009).

Since ACEs do not occur in isolation, but rather tend to co-occur among children. Those living with parents who have a mental illness tend to receive inadequate emotional care and easily be influenced by substance abuse and violence from parents, all of which increased the risks of cognitive impairment later in life.

Identifying how different ACEs exposure on depression and cognitive impairment in adulthood could help reduce the risks across the life course(Gehred et al., 2021). However, to date, the underlying mechanisms about ACEs exposure and mental health are still not well-known. One possible explanation is that prolonged stress caused by ACEs may lead to chronic activation of the hypothalamic-pituitary-adrenal (HPA) axis and then increase allostatic load and disrupt the regulation of neuroendocrine, immune and autonomic nervous (Kuhlman et al., 2018). Future studies are needed to enhance the understanding of the biological pathology of childhood adversities.

In this study, we did not observe the modification of certain sociodemographic variables on the association of ACEs and depression and cognitive impairment in later life. These results were consistent with several previous studies regardless of sex, household income status or socioeconomic situation (Houtepen et al., 2020; Lin et al., 2021). In contrast, some another studies reported that social support might mitigate the deleterious effects of childhood advertises on health outcomes (Cheong et al., 2017; Jaffee et al., 2017). These indicated that social resources, rather than individual-level characteristics, could be important modifiers to buffer the adverse effects of ACEs on adult health. Therefore, further explorations are needed to advance the current findings.

### Strengths and limitations

Our study has several strengths. Firstly, this was one of the few studies in a developing country to investigate a comprehensive range of ACE indicators and subsequent depression and cognitive impairment by a nationwide representative sample, which provided important implications that critical and persisting influence of childhood advertises might have across the life course. Secondly, we assessed the modification of the association by several sociodemographic characteristics, which built upon existing literature. Thirdly, the findings indicated that certain predictive factors such as physical abuse, household mental illness and domestic violence, which might help policy-makers or researchers to make intervention strategies to address mental health later in life. Lastly, the robustness of the association of accumulated ACEs and specific indicators of ACE with subsequent outcomes serves as a basis for creating a public health program to promote mental health across the life course.

Several limitations have been acknowledged. Firstly, the causal relationship is difficult to draw due to the cross-sectional design. Secondly, the ACE indicators were measured based on self-report, which might lead to recall bias. Thirdly, the ACE indicator of sexual abuse could not be evaluated due to the data was not available, despite a wide range of indicators being analyzed in our study. Lastly, the information on other potential confounders, such as physical activity, dietary nutrition, and medical treatment or assistive devices that mitigate the effect of mental illness were not collected. For all these limitations mentioned above, the findings should be interpreted with caution.

## Conclusions

In this nationwide population-based study, a dose-response association was observed between the accumulated ACEs and increased risks of subsequent depression and cognitive impairment among Chinese adults. The current findings indicate that sociodemographic characteristics of age, sex, educational level and family’s financial status during childhood did not modify such association. Considering China’s rapidly aging population and high prevalence of mental disorders, our study provides important public health implications for reducing childhood adversities and improving overall health and well-being in later life.

## Data Availability

All data produced in the present study are available upon reasonable request to the authors

## Authors’ contribution

Concept and design : WS; Acquisition, analysis, or interpretation of data: TZ and WS; Drafting of the manuscript: TZ and WS; Critical revision of the manuscript : LK, CJ, WS; Statistical analysis: WS; Supervision: WS

## Acknowledgement

We thank the Peking National Center for Economic Research for providing the CHARLS data. We are grateful to all the participants and on-site researchers in the project.

